# Vitamin D deficiency and its association with iron deficiency in African children

**DOI:** 10.1101/2021.07.14.21260499

**Authors:** Reagan M Mogire, John Muthii Muriuki, Alireza Morovat, Alexander J Mentzer, Emily L Webb, Wandia Kimita, Francis M Ndungu, Alex W Macharia, Clare L. Cutland, Sodiomon B Sirima, Amidou Diarra, Alfred B Tiono, Swaib A Lule, Shabir A Madhi, Andrew M. Prentice, Philip Bejon, John M Pettifor, Alison M Elliott, Adebowale Adeyemo, Thomas N Williams, Sarah H Atkinson

## Abstract

Vitamin D regulates the master iron hormone hepcidin, and iron in turn alters vitamin D metabolism. Although vitamin D and iron deficiency are highly prevalent globally little is known about their interactions in Africa. To evaluate associations between vitamin D and iron status we measured markers of iron status, inflammation, malaria parasitemia and 25-hydroxyvitamin D (25(OH)D) concentrations in 4509 children aged 0.3 months to 8 years living in Kenya, Uganda, Burkina Faso, The Gambia and South Africa. Prevalence of iron deficiency was 35.1%, and prevalence of vitamin D deficiency was 0.6% and 7.8% as defined by 25(OH)D concentrations of <30 nmol/L and <50 nmol/L respectively. Children with 25(OH)D concentrations of <50 nmol/L had a 98% increased risk of iron deficiency (OR 1.98 [95% CI 1.52, 2.58]) compared to those with 25(OH)D concentrations >75 nmol/L. 25(OH)D concentrations variably influenced individual markers of iron status. Inflammation interacted with 25(OH)D concentrations to predict ferritin levels. The link between vitamin D and iron status should be considered in strategies to manage these nutrient deficiencies in African children.

## 1. Introduction

Vitamin D and iron deficiency are two of the most common nutrient deficiencies worldwide [1,2]. Approximately 23% and 52% of children living in Africa are estimated to have vitamin D and iron deficiency, respectively [3,4]. *In vitro* and animal studies point to a complex interplay between vitamin D and iron metabolism (illustrated in Figure 1). Higher vitamin D status may improve iron status by decreasing levels of hepcidin, the principal iron-regulatory hormone, through the binding of the 1,25-dihydroxyvitamin D (1,25(OH)_2_D)–vitamin D receptor complex to the vitamin D response element (VDRE) on the hepcidin gene (*HAMP*) thus inhibiting its transcription [5], and by suppressing proinflammatory cytokines (IL6 and IL1B) [6], thus allowing iron absorption. Conversely, iron deficiency may also cause vitamin D deficiency by decreasing the activity of heme-containing vitamin D-activating enzymes such as 25- and 1α-hydroxylase, as demonstrated in rats [7], or by increasing fibroblast growth factor 23 (FGF23) [8], which suppresses the 1*α*-hydroxylation of vitamin D [9].

**Figure 1.**
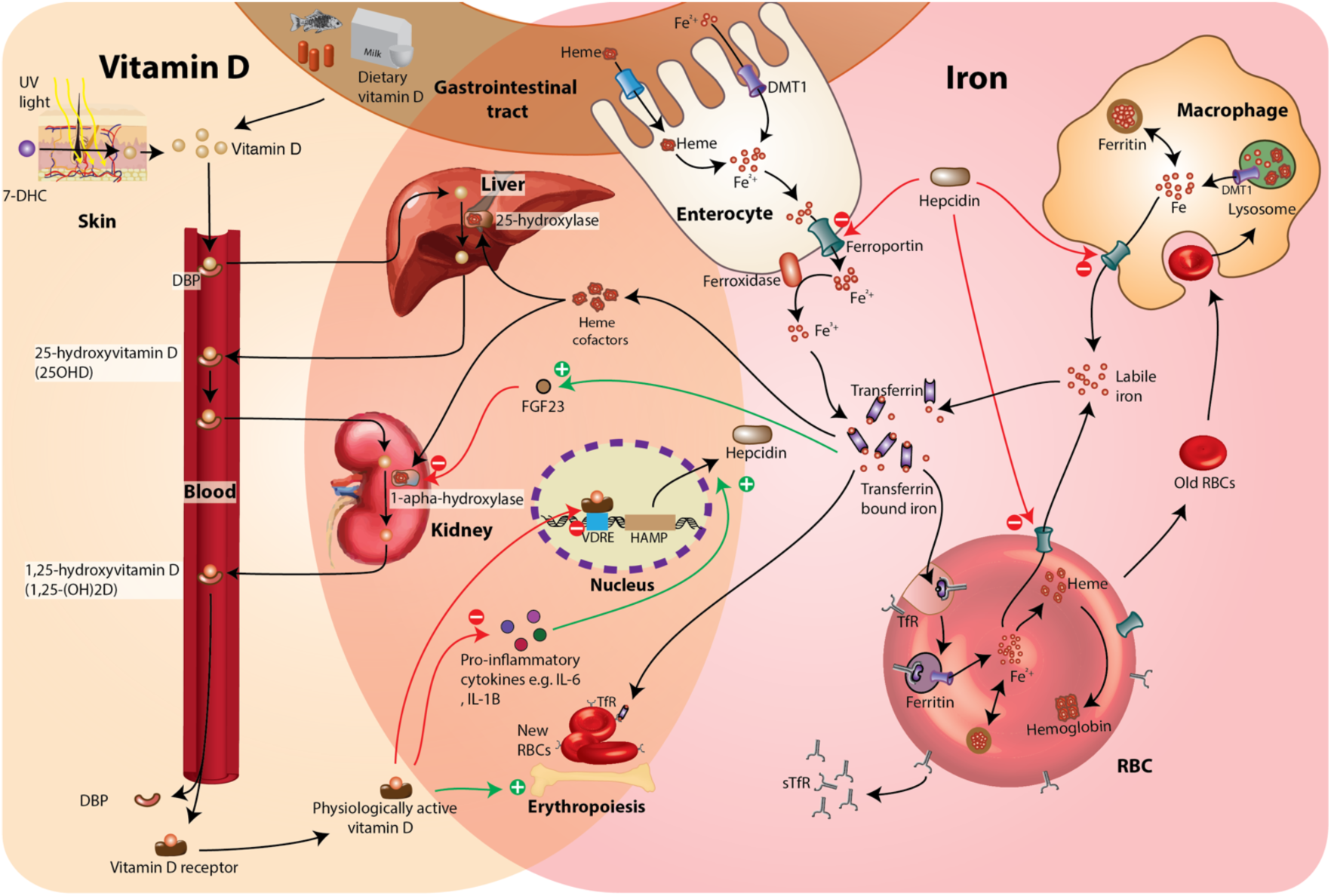
How the metabolism of vitamin D and iron is interlinked from *in vitro* and animal studies. 1,25(OH)_2_D may influence iron status by reducing hepcidin levels through directly binding to the vitamin D response element (VDRE) in the promoter region of the hepcidin gene (*HAMP*), decreasing pro-inflammatory cytokines (e.g. IL6, IL1B) and support erythropoiesis [5,6,13,14].Low iron status may also influence vitamin D status by decreasing the activity of vitamin D activation enzymes (25- and 1*α*-hydroxylase) [7] and increasing FGF23 [8]. High levels of FGF23 suppress 1*α*-hydroxylase activity thus reducing 1,25(OH)_2_D concentrations [9,15]. Abbreviations: sTfR, soluble transferrin receptor; DMT1, divalent metal transporter 1; DBP vitamin D binding protein; Cp, ceruloplasmin; FGF23, fibroblast growth factor 23; RBC, red blood cell; 7-DHC, 7-dehydrocholesterol.

Studies investigating the interactions between vitamin D and iron in humans have largely been in European adult populations and have reported mixed findings [10]. There are few studies in young children, a population with rapid growth requirements, or in Africa where malaria and other infectious diseases are also highly prevalent [11]. A single study in Kenyan preschool children (n = 500) found that 25-hydroxyvitamin (25(OH)D) concentrations were positively correlated with hemoglobin levels but did not measure markers of iron status [12]. The current study aimed to investigate associations between vitamin D and iron status in 4509 young children living in five countries across the African continent.

## 2. Materials and Methods

### Study population

This study included cross-sectional data from young children living in Kenya, Uganda, Burkina Faso, The Gambia and South Africa, as described below. A flowchart of study participant selection is presented in Figure S1.

*Kilifi, Kenya (0*.*3 months–8 years)*: This is an ongoing community-based cohort aimed at evaluating immunity to malaria in children living in Kilifi, Kenya [16]. Children are followed up from birth up to a maximum of 13 years with annual cross-sectional bleeds and weekly monitoring for malaria. 25(OH)D, iron and inflammatory biomarkers, anthropometry, and malaria parasitemia were measured from a single cross-sectional bleed based on the availability of archived plasma samples.

*Entebbe, Uganda (1–5 years):* The Entebbe Mother and Baby Study (EMaBS) is a prospective birth cohort study that was designed as a randomized controlled trial aimed at evaluating the effects of helminths and anthelmintic treatment on immunological and disease outcomes [17]. Blood samples were collected at birth and at subsequent birthdays up to five years of age. 25(OH)D, iron and inflammatory biomarkers, anthropometry, and malaria parasitemia were measured from a single annual visit based on the availability of stored samples.

*Banfora, Burkina Faso (6 months–2*.*5 years):* The VAC050 ME-TRAP Malaria Vaccine Trial was a clinical trial aimed at testing the effectiveness, safety and immunogenicity of a malaria vaccine in infants aged between six and 17 months [18]. 25(OH)D, iron and inflammatory biomarkers, anthropometry, and malaria parasitemia were measured at a single time point.

*West Kiang, The Gambia (2–6 years):* This study included children aged between two and six years recruited from rural villages in the West Kiang district in The Gambia before the malaria season as previously described [19]. Biomarkers, anthropometry and malaria parasitemia were measured during a single cross-sectional survey.

*Soweto, South Africa (4 months–2*.*8 years):* The Soweto Vaccine Response Study comprised of children who were participating in clinical vaccine trials. Samples for the current study were obtained from infants aged around 12 months. This cohort was not exposed to malaria and did not have anthropometric or hemoglobin measurements [20].

### Laboratory assays

25(OH)D concentrations, iron markers (ferritin, hepcidin, sTfR, transferrin, iron and hemoglobin) and inflammatory biomarkers (C-reactive protein [CRP] and α1-antichymotrypsin [ACT]) were assayed as previously described [4,21] and are summarized in Supplementary Methods 1. The assays showed satisfactory performances as monitored by both external quality assurance schemes (including UK National External Quality Assessment Scheme and DEQAS) and 4-hourly internal quality control assessments. The overall coefficient of variation for 25(OH)D measurements ranged from 2.8% to 7.9%. Over the duration of analyses, three sets of external quality assurance (DEQAS) data showed the 25(OH)D assay to have a mean (SD) bias of -2.7% (7.6) against the all-laboratory trended 25(OH)D values, and one of -0.4% (7.7) against the target values. Malaria parasitaemia was detected by blood film microscopy using the Giemsa staining technique.

### Definitions

Vitamin D status was defined using 25(OH)D cutoffs of <30 nmol/L, <50 nmol/L and 50–75 nmol/L [22,23]. Inflammation was defined as CRP levels >5 mg/L or ACT >0.6 g/L[24]. Iron deficiency was defined using WHO guidelines as plasma ferritin <12µg/L or <30µg/L in the presence of inflammation in children <5 years old, and as <15µg/L or <70µg/L in the presence of inflammation in children ≥5 years old [25]. Transferrin saturation (TSAT) was calculated as (iron in µmol/L/transferrin in g/L x 25.1) x 100 [26]. Anemia was defined as hemoglobin <11 g/dL in children aged <5 years, or hemoglobin <11.5 g/dL in children ≥5 years and iron deficiency anemia as the presence of both iron deficiency and anemia [27]. Malaria parasitaemia was defined as the presence of *Plasmodium* parasites in blood. Stunting was defined as height-for-age z-scores (HAZ) <-2 and underweight as weight-for-age (WAZ) <-2 according to 2006 WHO child growth standards [28].

### Statistical analyses

Statistical analyses were conducted using STATA 15.0 (StataCorp., College Station, TX). All biomarkers except transferrin and hemoglobin were natural log-transformed to normalize their distribution for regression analyses. Logistic and linear regression analyses were performed to evaluate the associations between vitamin D status (ln-25(OH)D concentrations and vitamin D status categories based on 25(OH)D concentrations <50, between 50–75, and >75 nmol/L) and iron deficiency, anemia and individual markers of iron status, where appropriate. Multivariable models were adjusted for study site, age, sex, season and inflammation. Stepwise backward regression analysis was used to determine suitable covariates to include in multivariable regression analyses. Regression models included only observations with values for all the variables included in the model. Since inflammation and malaria alter markers of iron status [4], we stratified by these variables in secondary multivariable regression analyses and tested for interactions between 25(OH)D concentrations and inflammation/malaria in predicting individual markers of iron status. Due to potential differences between study sites, we performed meta-analysis of site-specific multivariable regression estimates using the *metan* package in STATA.

## 3. Results

### 3.1 Characteristics of study participants

The study included 1361 Kenyan, 1301 Ugandan, 329 Burkinabe, 629 Gambian and 889 South African children. The characteristics of study participants are presented in Table 1. The children had a median age of 23.9 months (IQR 12.3, 35.9) and 49.1% were girls. Prevalence of stunting, underweight, inflammation and malaria parasitemia were high and varied by country. Overall median 25(OH)D concentrations were 77.6 (IQR 63.6, 94.2) nmol/L (Table 1). Prevalence of vitamin D deficiency was 0.6% and 7.8% using 25(OH)D cutoffs of <30 nmol/L and <50 nmol/L respectively, while 35.4% of children had concentrations between 50–75 nmol/L, as previously reported [21]. Since few children had 25(OH)D concentrations <30 nmol/L, only the <50 nmol/L cutoff was used for further analyses. Prevalence of iron deficiency, iron deficiency anemia and anemia was 35.1%, 23.0% and 61.6%, respectively, as previously reported [4]. South African children had the highest prevalence of low vitamin D status (25(OH)D <50 nmol/L) and iron deficiency at 13.5% and 42.0%, respectively.

**Table 1.** Characteristics of study participants

| Participant characteristics | Overall | Kenya | Uganda | Burkina Faso | The Gambia | South Africa<sup>8</sup> |
| --- | --- | --- | --- | --- | --- | --- |
| Total participants, *n*/total (%) | 4509 | 1361/4509 (30.2%) | 1301/4509 (28.9%) | 329/4509 (7.3%) | 629/4509 (13.9%) | 889/4509 (22.9%) |
| Median age in months (IQR) | 23.9 (12.3, 35.9) | 19.8 (12.7, 36.8) | 24.0 (23.9, 35.9) | 23.4 (19.7, 26.4) | 46.6 (35.2, 58.7) | 12.0 (11.9, 12.1) |
| Females, *n*/total (%) | 2216/4509 (49.1%) | 671/1361 (49.3%) | 641/1301 (49.3%) | 161/329 (48.9%) | 297/629 (47.2%) | 446/889 (50.2%) |
| Malaria parasitaemia<sup>1</sup>, *n*/total (%) | 445/3293 (13.5%) | 227/1082 (20.8%) | 89/1280 (6.9%) | 64/303 (21.1%) | 65/628 (10.4%) | n/a |
| Inflammation<sup>2</sup>, *n*/total (%) | 1019/4469 (22.8%) | 363/1344 (27.0%) | 306/1285 (23.8%) | 109/322 (33.9%) | 85/629 (13.5%) | 156/889 (17.6%) |
| Stunting, *n*/total (%)<sup>3</sup> | 581/2289 (25.4%) | 99/208 (47.6%) | 203/1282 (15.8%) | 103/307 (33.5%) | 176/492 (35.8%) | n/a |
| Underweight, *n*/total (%)<sup>4</sup> | 389/2487 (15.6%) | 102/389 (26.2%) | 103/1296 (8.0%) | 58/309 (18.8%) | 126/493 (25.6%) | n/a |
| **Vitamin D status** |  |  |  |  |  |  |
| Median 25(OH)D nmol/L (IQR) | 77.6 (63.6, 94.2) | 81.0 (66.3, 101.6) | 78.6 (65.1, 94.5) | 78.4 (64.5, 91.3) | 71.2 (59.1, 84.2) | 76.2 (60.6, 91.9) |
| 25(OH)D >75 nmol/l | 2485/4509 (55.1%) | 815/1361 (59.9%) | 756/1301 (58.1%) | 186/329 (56.5%) | 265/629 (42.1%) | 463/889 (52.1%) |
| 25(OH)D 50–75 nmol/l | 1674/4509 (37.1%) | 464/1361 (34.1%) | 479/1301 (36.8%) | 123/329 (37.4%) | 302/629 (48.0%) | 306/889 (34.4%) |
| 25(OH)D <50 nmol/l | 350/4509 (7.8%) | 82/1361 (6.0%) | 66/1301 (5.1%) | 20/329 (6.1%) | 62/629 (9.9%) | 120/889 (13.5%) |
| 25(OH)D <30 nmol/l | 28/4509 (0.6%) | 4/1361 (0.3%) | 5/1301 (0.4%) | 0 (0%) | 2/629 (0.3%) | 17/889 (1.9%) |
| **Iron status** |  |  |  |  |  |  |
| Iron deficiency<sup>5</sup>, *n*/total (%) | 1546/4399 (35.1%) | 491/1322 (37.1%) | 433/1240 (34.9%) | 115/319 (36.1%) | 134/629 (21.3%) | 373/889 (42.0%) |
| Iron deficiency anemia<sup>6</sup>, *n*/total (%) | 661/2880 (23.0%) | 207/771 (26.9%) | 209/1182 (17.7%) | 96/304 (31.6%) | 107/623 (17.2%) | n/a |
| Anemia<sup>7</sup>, *n*/total (%) | 1829/2971 (61.6%) | 556/793 (70.1%) | 623/1241 (50.2%) | 274/314 (87.3%) | 376/623 (62.4%) | n/a |

Abbreviations: 25(OH)D, 25-hydroxyvitamin D; IQR, interquartile range; n/a, not available or for malaria no exposure; CRP, C-reactive protein; ACT: α1-antichymotrypsin; sTfR, soluble transferrin receptors; TSAT, transferrin saturation. <sup>1</sup>Malaria parasitaemia was defined as the presence of *Plasmodium* parasitaemia on blood film; <sup>2</sup>inflammation as CRP > 5 mg/L or ACT >0.6 g/L (ACT, but not CRP was available for The Gambia); <sup>3</sup>stunting as height-for-age Z score <-2 and <sup>4</sup>underweight as weight-for-age Z score < -2; <sup>5</sup>iron deficiency as either plasma ferritin <12 μg/L or <30 μg/L in the presence of inflammation in children <5 years old, or <15 μg/L or <70 μg/L in the presence of inflammation in children ≥ 5 years old; <sup>6</sup>iron deficiency anemia as the presence of both iron deficiency and anemia; <sup>7</sup>anemia as hemoglobin <11 g/dL in children aged <5 years, or hemoglobin <11.5 g/dL in children ≥ 5 years. <sup>8</sup>Anthropometric and hemoglobin measurements were not available for South African children and they were not exposed to malaria.

### 3.2 Vitamin D status is associated with iron deficiency

The prevalence of iron deficiency was 41.8% (95% CI 36.7, 47.1) in children with 25(OH)D concentrations of <50 nmol/L compared to 35.8% (95% CI 33.9. 37.2) in those with concentrations >75 nmol/L. Similarly, the prevalence of vitamin D deficiency was higher in iron deficient children compared to those who were iron replete (Figure S2). In overall meta-analyses, 25(OH)D concentrations <50 nmol/L were associated with a 98% increased risk of iron deficiency (OR 1.98 [95% CI 1.52, 2.58]) compared to 25(OH)D concentrations >75 nmol/L (Figure 2, Table S1). We observed marked heterogeneity between the study countries for some of the meta-analyses (I^2^ = 68.7% to 90.4%, Figure 2). 25(OH)D concentrations <50 nmol/L were associated with reduced hemoglobin levels in overall meta-analyses (Figure 2). We observed increased odds of IDA (OR 1.42 [95% CI 0.96, 2.10]) and anemia (OR 1.27 [95% CI 0.90, 1.79]) in children with low vitamin D status (25(OH)D <50 nmol/L) but these associations were not statistically significant (Figure 2).

**Figure 2.**
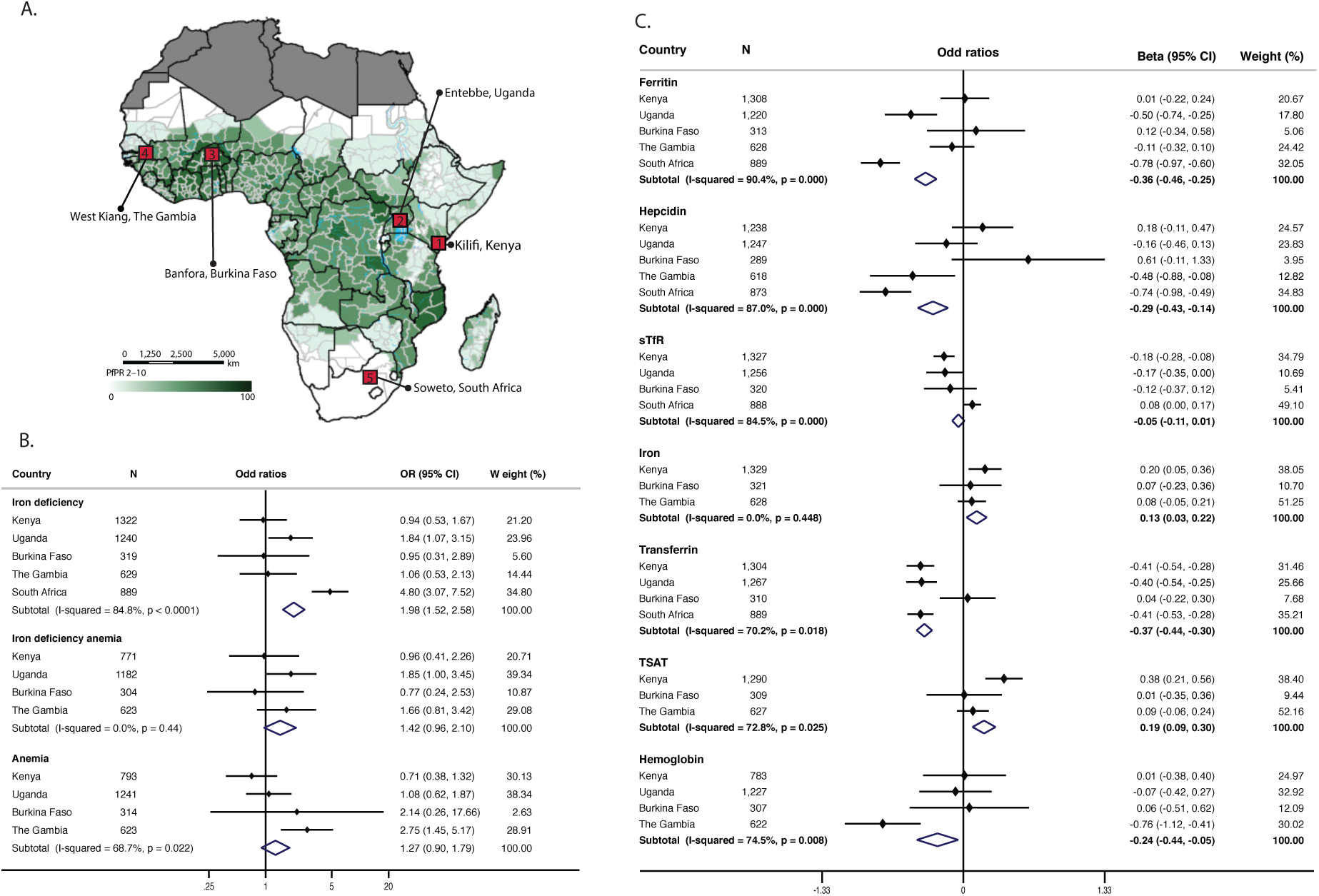
Map of Africa showing study sites (A), meta-analyses of associations between low vitamin D status (25(OH)D <50 nmol/L) and iron deficiency, iron deficiency anemia and anemia by study site (B) and meta-analyses of associations between low vitamin D status (25(OH)D <50 nmol/L) and individual markers of iron status (C). Abbreviations: sTfR, soluble transferrin receptors; TSAT, transferrin saturation. The map colors represent the predicted posterior predictions of age-standardized *P. falciparum* prevalence (PfPR2–10) as previously published by Snow et al [11]. Regression estimates were obtained from multivariable logistic regression analyses evaluating the effect of vitamin D status (25(OH)D <50 nmol/L against >75 nmol/L) on iron deficiency, iron deficiency anemia and anemia and on individual markers of iron status. Regression models were adjusted for age, sex, season, and inflammation. Meta-analysis of site-specific odd ratios was performed using *metan* package in STATA. Estimates for the meta-analyses for analyses using vitamin D status defined by 25(OH)D levels of 50-75 nmol/L are presented in Table S1.

### 3.3 Vitamin D status influences individual markers of iron status

Individual markers of iron status differed by country and vitamin D status (Tables S2 and S3, respectively). In overall meta-analyses children with 25(OH)D concentrations <50 nmol/L had reduced ferritin, hepcidin and hemoglobin levels compared to those with 25(OH)D concentrations >75 nmol/L (adj. beta -0.36 [95% CI -0.46, -0.025], -0.29 [95% CI - 0.43, -0.14], -0.24 [95% CI -0.44, -0.05], respectively) in keeping with increased risk of iron deficiency (Figure 2). However, other measures of iron status indicated a more complex picture, for example transferrin levels were lower, and iron levels and TSAT were higher, in children with 25(OH)D concentrations <50 nmol/L suggesting improved iron status (Figure 2). Similar results were obtained in pooled regression analyses (Table S3). 25(OH)D concentrations were highly correlated with hemoglobin levels and all iron markers except hepcidin (Figure S3). In pooled linear regression analyses controlling for age, sex, season, inflammation and study site, 25(OH)D concentrations were positively associated with ferritin, hepcidin, sTfR, and transferrin levels, and inversely associated with iron concentrations and TSAT (Figure S4).

### 3.4 ffects of inflammation and malaria on vitamin D and iron status

Since inflammation alters measures of iron status and differs by vitamin D status [21], we further stratified regression analyses by the presence of inflammation and looked for interactions between inflammation and 25(OH)D concentrations in predicting iron status. We observed that 25(OH)D concentrations were highly associated with measures of iron status in the absence of inflammation in multivariable linear regression analyses (Figure S4). We also found an interaction between inflammation and 25(OH)D concentrations in predicting ferritin levels, so that in children with inflammation a unit change in 25(OH)D concentrations was associated with a smaller unit change in ferritin levels than in children without inflammation (Table S4). Observed associations between 25(OH)D concentrations and markers of iron status persisted after excluding children with malaria (Figure S5) and we found little evidence of an interaction between malaria and 25(OH)D concentrations in predicting markers of iron status (Table S4).

## 4. Discussion

Nutritional deficiencies, including iron and vitamin D deficiency are highly prevalent among young children and little is known about their interactions in Africa where infectious diseases such as malaria are also prevalent. In the current study including 4509 children living in Africa, median 25(OH)D concentration was 77.0 nmol/L which was comparable to average concentrations in a meta-analysis of previous studies of young children in Africa (70.7 nmol/L) [21]. Low vitamin D status (25(OH)D concentrations <50 nmol/L) was associated with a 98% increased risk of iron deficiency and iron deficient children similarly had a higher prevalence of low vitamin D status. In keeping with increased iron deficiency low vitamin D status was associated with reduced levels of ferritin, hepcidin and hemoglobin. We found marked heterogeneity in the study site estimates, with the strongest association between low vitamin D status and iron deficiency observed in South Africa. 25(OH)D concentrations interacted with inflammation, so that in children with inflammation higher 25(OH)D concentrations predicted lower ferritin concentrations than in those without inflammation. We found limited evidence of increased risk of iron deficiency anemia and anemia in children with low vitamin D status in overall meta-analyses.

Low vitamin D status (25(OH)D <50 nmol/L) was associated with an increased risk of iron deficiency and low ferritin levels in young African children in this study. Our findings are consistent with a study from South Korea which reported that young children with iron deficiency were four times more likely to have low vitamin D status (25(OH)D <50 and <75 nmol/L) compared to those who were iron replete (OR 4.12, 95% CI 1.67,10.17), although 25(OH)D levels were not correlated with ferritin levels (Table 2, [29]). Similarly, a nationally representative survey of 2526 children and young adults aged between 10 to 20 years from South Korea showed that vitamin D deficiency (25(OH)D concentrations m<37.5 nmol/L) was associated with a 94% increased odds of iron deficiency (OR 1.94 [95% CI, 1.27–2.97]) [30]. However, vitamin D status was not associated with ferritin levels in 22-month-old Asian infants in the UK [31], children attending hospital in northern India [32] or in children diagnosed with IDA in South Korea [33]. Mechanistic studies have shown that vitamin D may improve iron deficiency by reducing hepcidin levels through inhibiting its transcription or suppressing pro-inflammatory cytokines, thus allowing iron absorption [6].

**Table 2.** Summary of previous observational studies that evaluated the association between vitamin D status and markers of iron status and hemoglobin levels in children (<13 years old)

| Author, year | Sample size (*n*) | Country | Age | Population | Definition of vitamin D status | Markers investigated | Findings |
| --- | --- | --- | --- | --- | --- | --- | --- |
| **Iron markers** |  |  |  |  |  |  |  |
| Grindulis 1986 [31] | 145 | UK | 22 months | Asian children | VDD: <25 nmol/L | Serum iron, transferrin, ferritin and hemoglobin | Children with VDD had lower concentrations of hemoglobin and serum iron. No differences were observed with transferrin and ferritin levels. |
| Yoon 2012 [33] | 79 | South Korea | Median 1.8 (IQR 0.3–13) years | Children diagnosed with IDA | VDS: ≥75 nmol/L<br>VDI: 30–75 nmol/L<br>VDD: <30 nmol/L | Ferritin, serum iron and hemoglobin | No difference in ferritin, serum iron or hemoglobin levels in patients with VDS compared to those who had VDD or VDI. No correlation between vitamin D status and severity of anemia. |
| Jin 2013 [29] | 102 | South Korea | Range 3–24 months | Children attending hospital | VDS: ≥75 nmol/L<br>VDI: 50–75 nmol/L<br>VDD: <50 nmol/L | Ferritin and iron | Children with ID were four times more likely to develop VDI and VDD compared to those who were iron replete. 25(OH)D levels were not correlated with ferritin or iron levels. |
| Sharma 2015 [32] | 263 | North India | Range 3 months–12 years | Children attending outpatients | VDS: ≥75 nmol/L<br>VDI: 30–75 nmol/L<br>VDD: <30 nmol/L | Ferritin, serum iron and hemoglobin | The proportion of children with IDA was 66%, 49% and 25% in the VDD, VDI group and VDS groups, respectively. 25(OH)D levels were positively correlated with hemoglobin levels but not with ferritin or iron levels. |
| Cihan 2018 [34] | 117 | Turkey | 6 months–5 years | Children attending outpatients | VDS: ≥50 nmol/L<br>VDI: 30–50 nmol/L<br>VDD: <30 nmol/L | Serum iron, ferritin and hemoglobin | Maternal and child VDD was associated with ID / IDA in children. Children with ID/IDA had lower 25(OH)D levels than those without ID/IDA. Maternal/child 25(OH)D levels were positively correlated with hemoglobin levels in children. |
| Chowdhury 2019 [35] | 1000 | India | Range 0.5–2.5 years | Community-based children | VDD: <25 nmol/L | Hemoglobin and sTfR | Children with VDD were more likely to have anemia independently of ID. Hemoglobin levels were positively associated with 25(OH)D levels. Association between 25(OH)D and sTfR was not investigated. |
| **Hemoglobin only** |  |  |  |  |  |  |  |
| Abdul-Razzak 2011 [35] | 203 | Jordan | Range 0.5–3 years | Infants and children attending primary care | VDS: ≥75 nmol/L<br>VDI: <75 nmol/L<br>VDD: <50 nmol/L | Hemoglobin | No difference in mean hemoglobin levels between vitamin D status categories (VDD, VDI and VDS). |
| Kang 2015 [36] | 70 | South Korea | Range 4–24 months | Mothers and their infants attending hospital | VDS: ≥75 nmol/L<br>VDI: 50–75 nmol/L<br>VDD: <50 nmol/L | Hemoglobin | Mothers and infants with anemia had lower 25(OH)D levels compared to those without anemia. |
| Houghton 2019 [37] | 120 | India | 12–23 months | Community-based children | VDD: <50 nmol/L | Hemoglobin | No association between 25(OH)D levels and anemia. |
| Houghton 2019 [12] | 500 | Kenya | 3–5 years | Preschool children | VDD: <50 nmol/L | Hemoglobin | 25(OH)D levels were positively correlated with hemoglobin levels. |

Abbreviations: 25(OH)D, 25-hydroxyvitamin D; VDS, vitamin D sufficiency; VDI, vitamin D insufficiency; VDD, vitamin D deficiency; ID, iron deficiency; IDA, iron deficiency anemia; IQR, interquartile range.

In the current study, hepcidin levels were lower in children with low vitamin D status (25(OH)D <50 nmol/L) compared to those with 25(OH)D concentrations >75 nmol/L. 25(OH)D concentrations were also inversely associated with hepcidin levels in multivariable models. To the best of our knowledge, no previous studies have investigated the association between vitamin D and hepcidin levels in young children, although studies in older patients with rheumatoid arthritis and inflammatory bowel disease have reported mixed findings [38,39]. *In vitro* and animal studies indicate that vitamin D can decrease hepcidin levels through a number of pathways including inhibiting transcription of the hepcidin gene (HAMP), suppressing the production of pro-inflammatory cytokines IL-6 and IL-1B, or by activating the JAK-STAT3 pathway (Figure 1) [5,6,13]. The converse finding of lower hepcidin levels in vitamin D deficient children in our study is likely due to iron deficiency exerting a powerful inhibitory effect on hepcidin expression in African children [40].

Low vitamin D status (25(OH)D <50 nmol/L) was associated with increased iron concentrations and TSAT and decreased transferrin levels compared to 25(OH)D concentrations >75 nmol/L. In contrast to our study, an earlier study including 22-month-old Asian infants reported that vitamin D deficiency (25(OH)D <25 nmol/L) was associated with lower serum iron concentrations, but not transferrin levels [31]. Two studies that included children from South Korea and North India reported no association between vitamin D status and iron concentrations (Table 2, [32,33]). A single study investigating the association between TSAT and 25(OH)D concentrations found that higher 25(OH)D concentrations were associated with increased TSAT in healthy adolescent Saudi boys, although this was not observed in girls or in pooled analyses. We found that sTfR levels did not differ by vitamin D status in our overall meta-analysis. In contrast to our study, vitamin D deficiency was associated with increased sTfR levels in observational studies involving children and adolescents in Germany [41] and in Polish athletes [42]. In the current study, it is possible that the lower hepcidin levels observed in vitamin D deficient children may have led to increased iron absorption [43] resulting in higher iron concentrations and TSAT. Children in this study and particularly those living in sub-Saharan Africa would also be likely to have had more infections including malaria and nutritional deficiencies which might explain differences in findings.

In the current study, low vitamin D status (25(OH)D <50 nmol/L) was associated with reduced hemoglobin levels, although the association with anemia was not statistically significant. In agreement with our study, studies in children aged 3–5 years in Kenya, 22 months in the UK, and between 0.5–2.5 years in India, reported a positive association between vitamin D status and hemoglobin levels (Table 2, [12,31,44]). However, a study involving healthy children living in Jordan (n= 203, aged 0.5 – 3 years) did not find an association between vitamin D status and hemoglobin levels or anemia [35]. A systematic review and meta-analysis of seven observational studies with 5183 participants showed that vitamin D deficiency was associated with 84% higher odds of anemia (OR 2.25, 95% CI 1.47-3.44) [45]. Anemia may be caused by many factors in addition to iron deficiency in African children, including malaria, undernutrition, hemoglobinopathies and sickle cell disease, which may explain why we did not observe an association between vitamin D status and anemia in our study.

Conversely, iron status is also likely to have influenced vitamin D status and we observed a higher prevalence of low vitamin D status (25(OH)D <50 nmol/L) in children with iron deficiency. Since our study was observational, we were unable to ascertain the direction of causality in the association between vitamin D and iron status. Iron deficiency may lead to vitamin D deficiency via several pathways including by reducing the activity of heme-containing vitamin D activation enzymes such as 25-hydroxylase and 1-alpha-hydroxylase [7] or by reducing vitamin D absorption in the gastrointestinal tract through impairing epithelial function [46] (Figure 1). Iron deficiency has also been shown to increase levels of fibroblast growth factor 23 (FGF23) levels [47,48] which suppresses 1 *α*-hydroxylase activity [9,49]. A strong inverse correlation between FGF23 and hemoglobin mhas been suggested to link low iron status in the etiology of rickets in Gambian children [50]. In contrast, pregnant Kenyan women (n = 433) supplemented with iron from 13–23 weeks of gestation until 1 month postpartum had lower 25(OH)D concentrations than those who received a placebo, although no differences were observed in 25(OH)D concentrations in cord blood [48].

The prevalence of inflammation and malaria parasitemia was high among our study participants and both vitamin D and iron status have been associated with increased risk of malaria [51,52]. We found that associations between vitamin D and iron status persisted after adjusting for inflammation and malaria in our analyses. Inflammation interacted with 25(OH)D concentrations in our study to predict ferritin levels, so that 25(OH)D concentrations most strongly predicted ferritin levels in children without inflammation. This may be one explanation for why some of our findings varied by country since malaria and inflammation were generally more highly prevalent in sub-Saharan African countries. The association between vitamin D status and iron deficiency was strongest in South African children who were not exposed to malaria and who generally had lower prevalence of inflammation. Other factors may have also contributed to heterogeneity in our findings, including variability in socio-cultural practices, nutrition, genetics (such as hemoglobinopathies or vitamin D binding protein variants) or seasonality in Africa.

Our study had a number of strengths. To the best of our knowledge, this is the first study to investigate the association between vitamin D status and iron deficiency in African children and the largest study in young children globally. Children living in sub-Saharan Africa have a high burden of iron deficiency, undernutrition and infectious diseases, which may explain why some of our findings differed from studies in other parts of the world. Additionally, we included a large number of community-based children (n = 4509) living in five different countries across Africa. We also measured a wide range of iron biomarkers which enabled us to evaluate how vitamin D status is associated with various aspects of iron status. Our study also had some limitations. The study was crosssectional hence we could not evaluate temporal changes in the observed associations or determine the direction of causality. There may have also been other unmeasured confounders that influenced our findings and may have accounted for the heterogeneity that we observed between study sites. We also only included young children in our study which may limit the generalizability of our findings to other age groups. Additionally, plasma iron levels and TSAT were not available for Ugandan and South African children and CRP measurements were not available for Gambian children. We also did not collect information that may have influenced vitamin D or iron status such as dietary intake, sunlight exposure or hookworm infections and did not measure 1,25(OH)_2_D, FGF23, vitamin D binding protein, parathyroid hormone, or calcium concentrations.

## 5. Conclusions

In conclusion, our findings suggest a complex association between vitamin D and iron status in young African children, with the direction of association varying depending on the iron status marker. Low vitamin D status (25(OH)D <50 nmol/L) was associated with iron deficiency as indicated by reduced ferritin, hepcidin and hemoglobin levels, although increased plasma iron and TSAT may suggest that these children may be more iron replete. The positive association between vitamin D and WHO-defined iron deficiency in this study suggest that both conditions should be considered in relevant public health strategies in Africa where the two nutrient deficiencies coexist. There is a need for further studies to understand the mechanisms of action of the associations between vitamin D and iron deficiency in African populations.

## Data Availability

The data and analyses scripts underlying this article are available in Harvard Dataverse at https://doi.org/10.7910/DVN/A78P8B and applications for data access can be made through the Kilifi Data Governance Committee (.

## Supplementary Materials

The following supporting information can be downloaded at: www.mdpi.com/xxx/s1, Supplementary Methods 1; Table S1: Effects of vitamin D status on iron deficiency, iron deficiency anemia and anemia in African children; Table S2: Geometric means of biomarkers by country; Table S3: Effect of vitamin D status on individual measures of iron status; Table S4: Interaction between 25(OH)D concentrations and inflammation and malaria in predicting measures of iron status; Figure S1: Flow chart of study participants; Figure S2: Prevalence of iron status by categories of vitamin D status (i) and prevalence of vitamin D status by categories of iron status (ii) in African children; Figure S3: Scatter and regression plots of markers of iron status and C-reactive protein against log 25(OH)D concentrations; Figure S4: Association between 25(OH)D concentrations and iron markers overall (dark grey) and in African children with and without inflammation (black and light grey lines respectively) Figure S5: Association between 25(OH)D concentrations and iron markers overall (blue) and in children with and without malaria in Africa (red and green lines, respectively).

## Author Contributions

RMM, JMM, AA, TNW and SHA conceived and designed the study. Data was obtained by all the authors and analysed by RMM, JMM, and SHA. RMM and SHA wrote the first draft of the manuscript. RMM, JMM, AJM, EMW, WK, AWM, FMN, CC, SBS, AD, ABT, SAL, AM, SAM, AMP, PB, JMP, AME, AA, TNW, SHA contributed to data interpretation, reviewed successive drafts and approved the final version of the manuscript. The sponsors played no role in the study design, data collection, data analysis, data interpretation or writing of the report

## Institutional Review Board Statement

Ethical approvals were granted by the Scientific Ethics Review Unit of the Kenya Medical Research Institute (KEMRI/SERU/CGMR-C/046/3257) in Kenya, the Uganda Virus Research Institute (GC/127/12/07/32) and Uganda National Council for Science and Technology (MV625) in Uganda, by Ministere de la Recherche Scientifique et de l’Innovation (reference 2014-12-151) in Burkina Faso, the Gambian Government and the Medical Research Council Ethics Review Committee in The Gambia (874/830), the University of Witwatersrand Human Research Ethics Committee (M130714) in South Africa and in the UK by the London School of Hygiene and Tropical Medicine (A340) and the Oxford Tropical Research Ethics Committees (39-12, 41-12, 42-14, and 1042-13).

## Informed Consent Statement

Informed written consent was obtained from all children’s parents or guardians before inclusion in the study.

## Data Availability Statement

The data and analyses scripts underlying this article are available in Harvard Dataverse at https://doi.org/10.7910/DVN/A78P8B and applications for data access can be made through the Kilifi Data Governance Committee.

## Acknowledgments

In this section, you can acknowledge any support given which is not covered by the author contribution or funding sections. This may include administrative and technical support, or donations in kind (e.g., materials used for experiments).

---

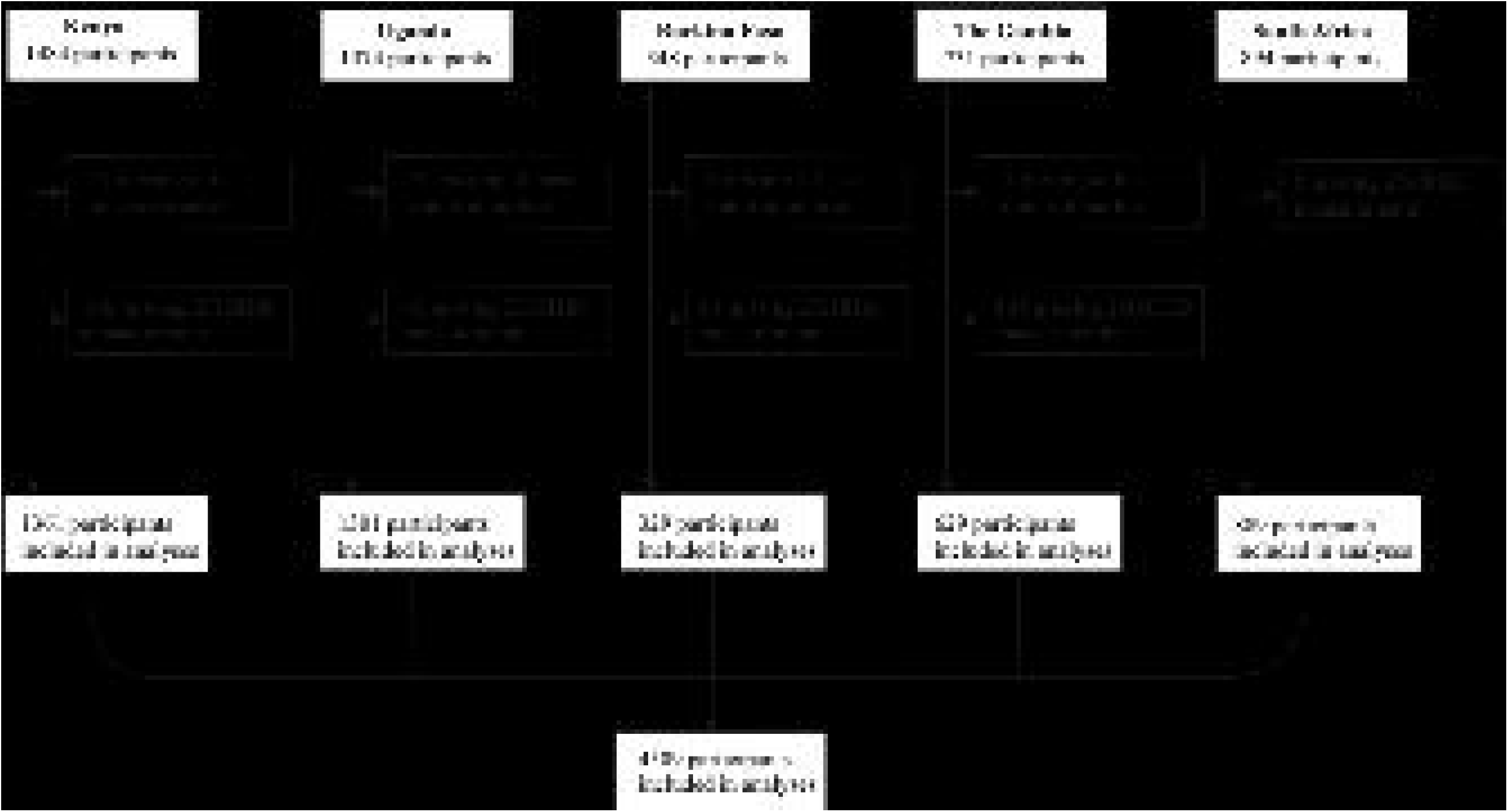

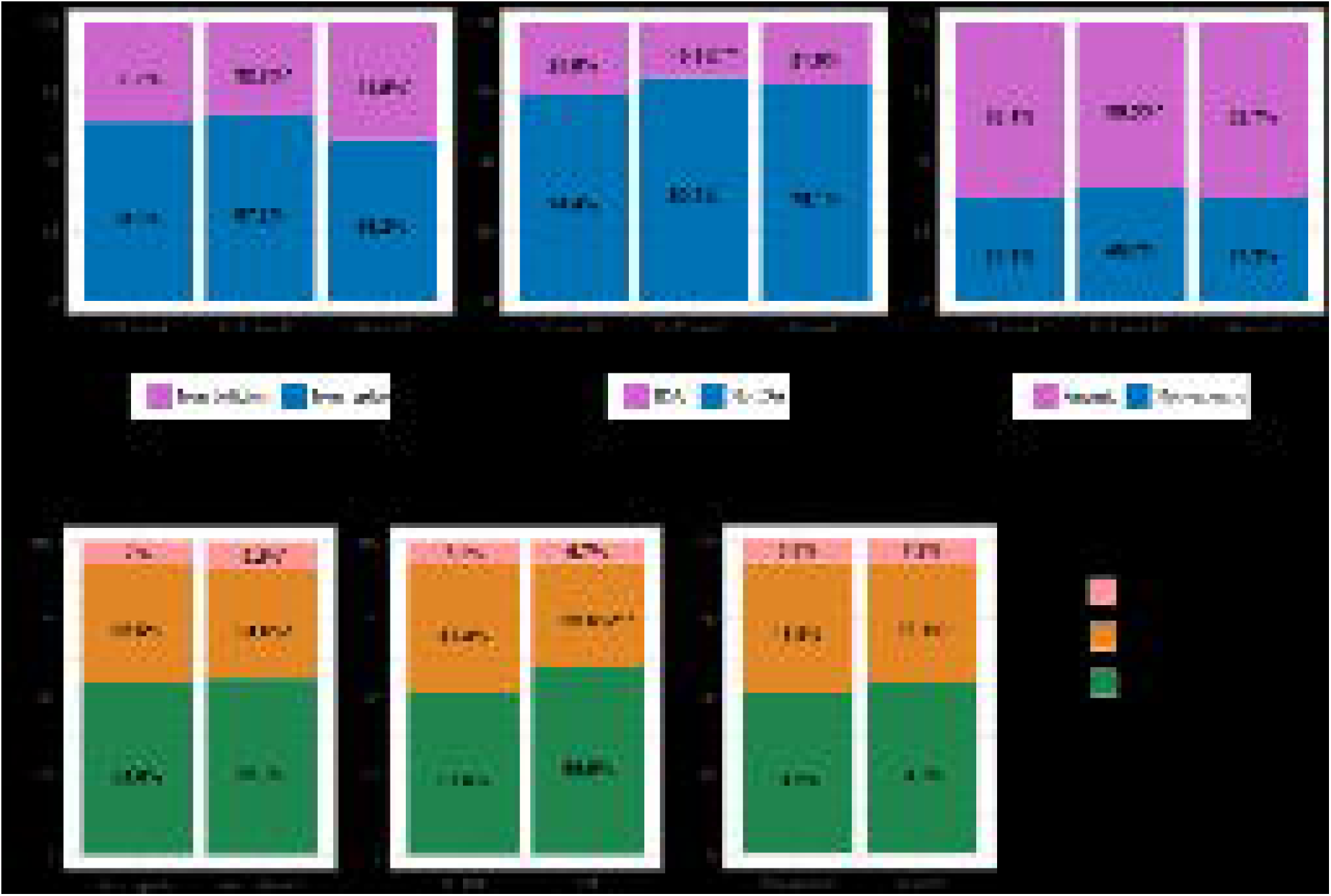

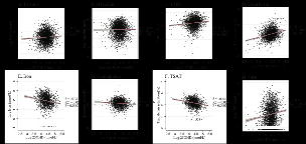

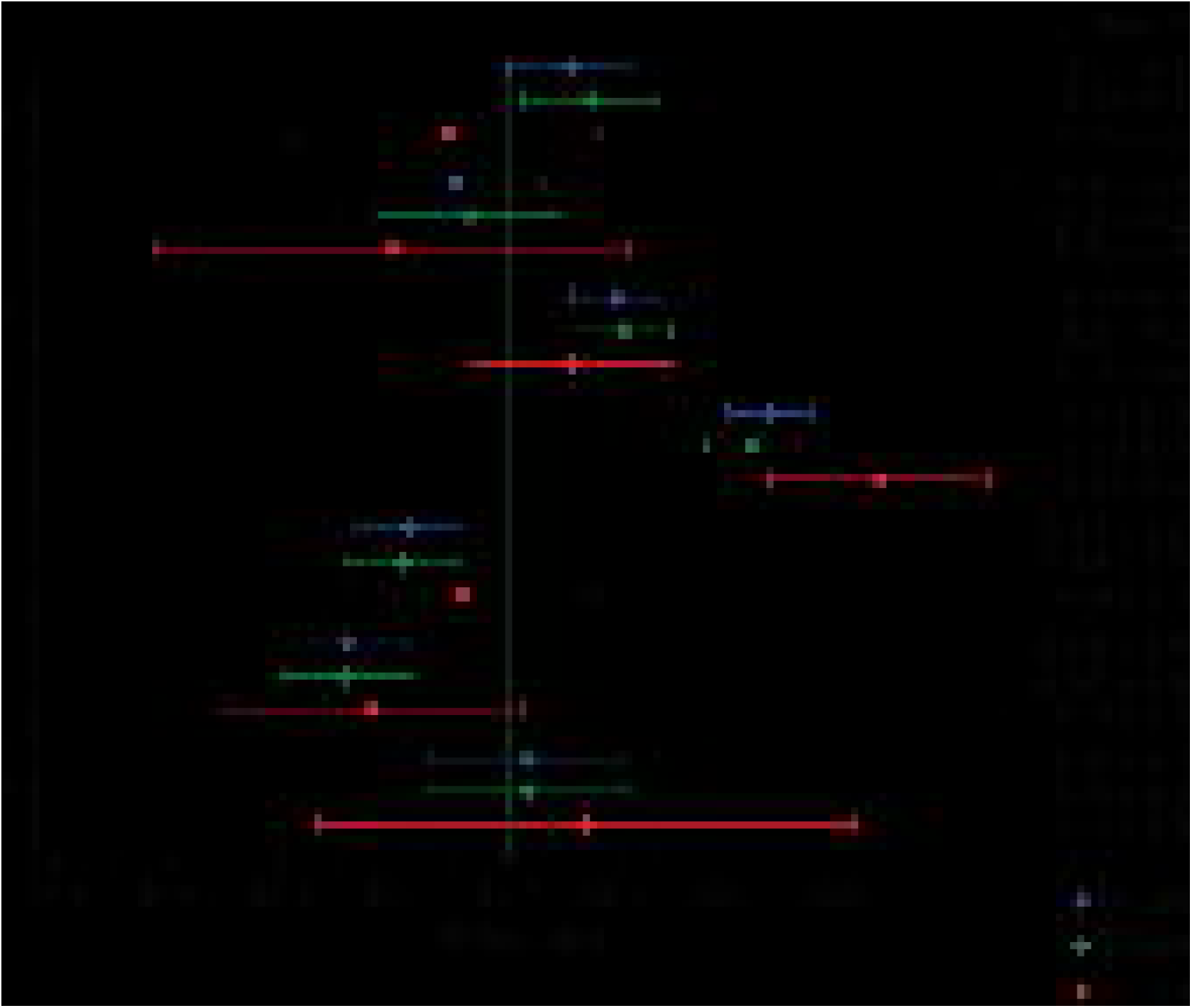

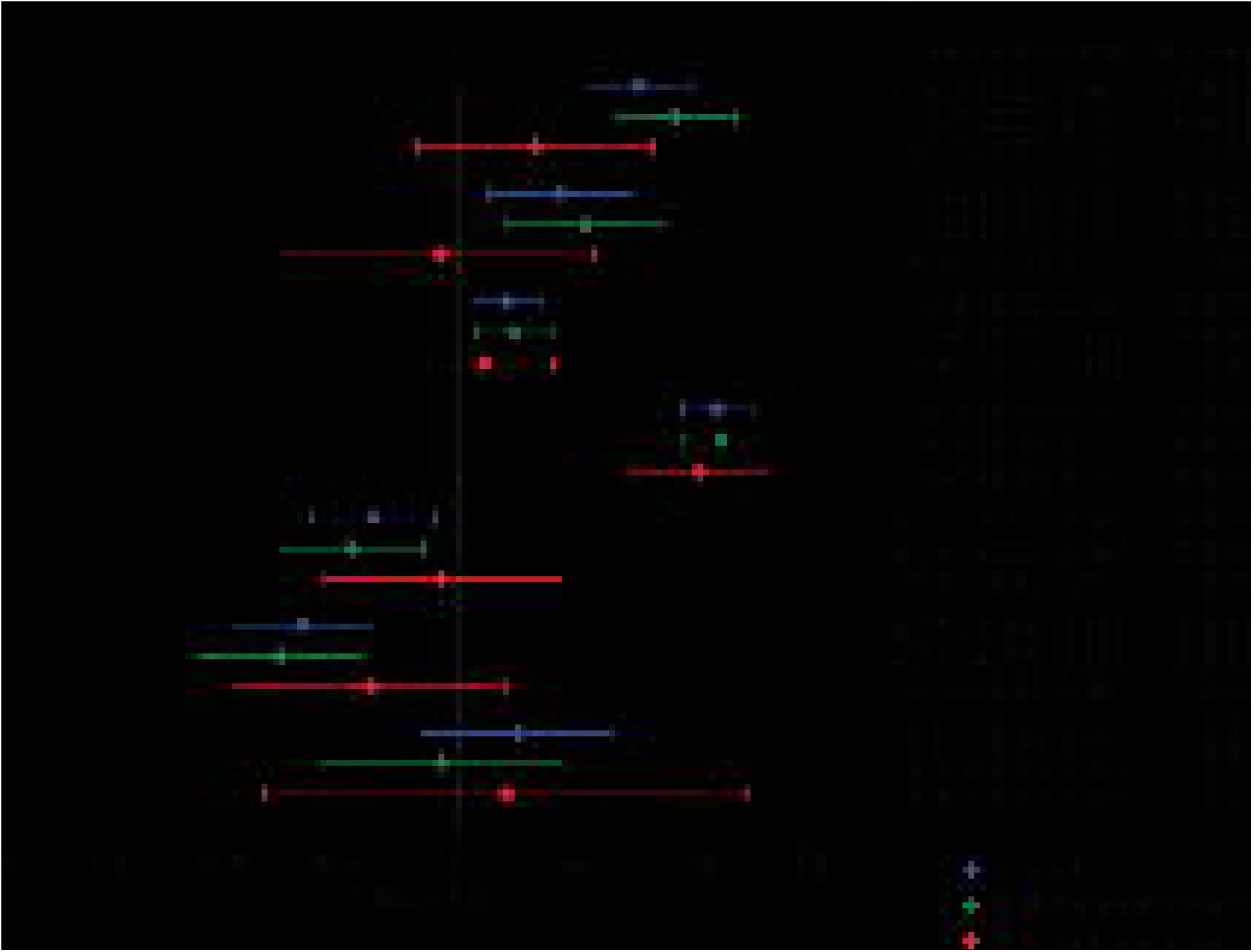

