## Supplementary Materials for "Vitamin D deficiency and its association with iron deficiency in African children"

**Supplementary Methods 1. Biochemical assays for vitamin D, iron and inflammatory biomarkers by study site**

| **Site** | **25(OH)D** | **Ferritin** | **Hepcidin** | **sTfR** | **Iron** | **Transferrin** | **CRP** | **ACT** |
| --- | --- | --- | --- | --- | --- | --- | --- | --- |
| **Kilifi, Kenya** | Chemiluminescent microparticle immunoassay (Abbott Architect, USA) | Microparticle Enzyme Immunoassay (Abbott Architect, USA) | DRG Hepcidin 25 [bioactive] high sensitive ELISA (DRG International, USA) | Human sTfR ELISA (BioVendor, Czech Republic) | MULTIGENT iron calorimetric assay, Abbott Architect, USA | Chemiluminescent Microparticle Immunoassay (Abbott Architect, USA) | MULTIGENT CRP Vario assay, (Abbott Architect, USA) | Not measured |
| **Banfora, Burkina Faso** | Chemiluminescent microparticle immunoassay (Abbott Architect, USA) | Microparticle Enzyme Immunoassay (Abbott Architect, USA) | DRG Hepcidin 25 [bioactive] high sensitive ELISA (DRG International, USA) | Human sTfR ELISA (BioVendor, Czech Republic) | MULTIGENT iron calorimetric assay, Abbott Architect, USA | Chemiluminescent Microparticle Immunoassay (Abbott Architect, USA) | MULTIGENT CRP Vario assay, (Abbott Architect, USA) | Not measured |
| **Entebbe, Uganda** | Chemiluminescent microparticle immunoassay (Abbott Architect, USA) | Microparticle Enzyme Immunoassay (Abbott Architect, USA) | DRG Hepcidin 25 [bioactive] high sensitive ELISA (DRG International, USA) | Human sTfR ELISA (BioVendor, Czech Republic) | Not measured | Chemiluminescent Microparticle Immunoassay (Abbott Architect, USA) | MULTIGENT CRP Vario assay, (Abbott Architect, USA) | Not measured |
| **Soweto, South Africa** | Chemiluminescent microparticle immunoassay (Abbott Architect, USA) | Microparticle Enzyme Immunoassay (Abbott Architect, USA) | DRG Hepcidin 25 [bioactive] high sensitive ELISA (DRG International, USA) | Human sTfR ELISA (BioVendor, Czech Republic) | Not measured | Chemiluminescent Microparticle Immunoassay (Abbott Architect, USA) | MULTIGENT CRP Vario assay, (Abbott Architect, USA) | Not measured |
| **West Kiang, The Gambia** | Chemiluminescent microparticle immunoassay (Abbott Architect, USA) | Microparticle Enzyme Immunoassay (Abbott Architect, USA) | Hepcidin-25 [human] Enzyme Immunoassay Kit (Bachem, Switzerland) | Quantikine sTfR ELISA kit, (R&D Systems, USA) | Ferrozine based photometry and colorimetry analyser (Hitachi 911, Tokyo, Japan) | Not measured | Not measured | Immunoturbidimetry Cobas Mira Plus Bioanalyser, Roche |
| Abbreviations: ACT: α1-antichymotrypsin; CRP: C-reactive protein; ELISA: enzyme-linked immunosorbent assay; and sTfR: soluble transferrin receptor. Hemoglobin levels for Ugandan children were adjusted for an altitude of greater than 1 km above sea level by subtracting 0.2 g/dL [52]. Iron measurements were not available for Ugandan and South African cohorts because their plasma samples were stored in ethylenediaminetetraacetic acid (EDTA) which chelates iron. Hepcidin values in Gambian samples were harmonized by first converting to the old DRG hepcidin assay values and thereafter to the new highly sensitive DRG hepcidin assay values [53]. | | | | | | | | |

**Table S1. Effects of vitamin D status on iron deficiency, iron deficiency anemia and anemia in African children**

|  |  | **Vitamin D status categories based on 25(OH)D levels** | | | | | | | | |
| --- | --- | --- | --- | --- | --- | --- | --- | --- | --- | --- |
|  |  | **>75 nmol/L (reference)** |  | **50–75 nmol/L** | | |  | **<50 nmol/L** | | |
| **Proportion, *n*/total (%)** |  | 2485/4509 (55.1%) |  | 1674/4509 (37.1%) | | |  | 350/4509 (7.8%) | | |
|  |  | ***n*** |  | ***n*** | **OR (95% CI)** | ***p*** |  | ***n*** | **OR (95% CI)** | ***p*** |
| **Iron deficiency**^1^ | Overall^4^  Kenya  Uganda  Burkina Faso  The Gambia  South Africa | 2425/4399  794/1322  725/1240  178/319  265/629  463/889 |  | 1632/4399  450/1322  452/1240  122/319  302/629  306/889 | 1.12 (0.97, 1.30)  0.86 (0.65, 1.13)  0.95 (0.73, 1.23)  1.12 (0.66, 1.90)  0.82 (0.54, 1.24)  2.45 (1.78, 3.37) | 0.13  0.27  0.68  0.67  0.35  <0.0001 |  | 342/4399  78/1322  63/1240  19/319  62/629  120/889 | 1.98 (1.52, 2.58)  0.94 (0.53, 1.67)  1.84 (1.07, 3.15)  0.95 (0.31, 2.89)  1.06 (0.53, 2.13)  4.80 (3.07, 7.52) | <0.0001  0.84  0.027  0.93  0.86  <0.0001 |
| **Iron deficiency anemia**^2^ | Overall^4^  Kenya  Uganda  Burkina Faso  The Gambia  South Africa | 1540/2880  420/771  690/1182  167/304  263/623  n/a |  | 1139/2880  293/771  430//1182  118/304  298/623  n/a | 0.94 (0.76, 1.15)  0.91 (0.61, 1.38)  0.91 (0.67, 1.25)  1.01 (0.57, 1.76)  0.98 (0.62, 1.57)  n/a | 0.52  0.68  0.57  0.98  0.96  n/a |  | 201/2880  58/771  62//1182  19/304  62/623  n/a | 1.42 (0.96, 2.10)  0.96 (0.41, 2.26)  1.85 (1.00, 3.45)  0.77 (0.24, 2.53)  1.66 (0.81, 3.42)  n/a | 0.075  0.93  0.052  0.67  0.17  n/a |
| **Anemia**^3^ | Overall^4^  Kenya  Uganda  Burkina Faso  The Gambia  South Africa | 1587/2971  430/793  719/1241  175/314  263/623  n/a |  | 1175/2971  301/793  457/1241  119/314  298/623  n/a | 1.03 (0.87, 1.23)  0.65 (0.45, 0.93)  1.08 (0.84, 1.39)  1.37 (0.65, 2.88)  1.35 (0.96, 1.91)  n/a | 0.73  0.019  0.55  0.41  0.086  – |  | 209/2971  65/793  65/1241  20/314  62/623  n/a | 1.27 (0.90, 1.79)  0.71 (0.38, 1.32)  1.08 (0.62, 1.87)  2.14 (0.26, 17.66)  2.75 (1.45, 5.17)  n/a | 0.17  0.28  0.79  0.48  0.002  – |
| Abbreviations: 25(OH)D, 25-hydroxyvitamin D; n/a, not available. Odds ratios and *p* values were obtained from multivariable logistic regression analyses adjusted for age, sex, season, inflammation and study site. 25(OH)D and CRP levels were ln-transformed to make them normally distributed. ^1^Iron deficiency was defined as either plasma ferritin <12µg/L or <30µg/L in the presence of inflammation in children <5 years old, or <15µg/L or <70µg/L in the presence of inflammation in children ≥5 years old; ^2^iron deficiency anemia as the presence of both iron deficiency and anemia; ^3^anemia was defined as hemoglobin <11 g/dL in children aged <5 years, or hemoglobin <11.5 g/dL in children ≥5 years; inflammation as CRP > 5 mg/L or ACT >0.6 g/L (ACT, but not CRP was available for The Gambia). The overall estimate was obtained by performing a meta-analysis of study site estimates using *metan* in STATA. | | | | | | | | | | |

**Table S2. Geometric means of biomarkers by country**

| **Biomarker means** |  | **Overall** |  |  | **Kenya** |  |  | **Uganda** |  |  | **Burkina Faso** |  |  | **The Gambia** |  |  | **South Africa**^#^ |
| --- | --- | --- | --- | --- | --- | --- | --- | --- | --- | --- | --- | --- | --- | --- | --- | --- | --- |
|  | ***n*** | **Mean (95% CI)** |  | ***n*** | **Mean (95% CI)** |  | ***n*** | **Mean (95% CI)** |  | ***n*** | **Mean (95% CI)** |  | ***n*** | **Mean (95% CI)** |  | **n** | **Mean (95% CI)** |
| 25(OH)D (nmol/L) | 4509 | 77.0 (76.3, 77.7) |  | 1361 | 81.9 (80.4, 83.3) |  | 1301 | 78.3 (77.0, 79.5) |  | 329 | 77.3 (75.0, 79.6) |  | 629 | 70.4 (69.0, 71.9) |  | 889 | 72.8 (71.2, 74.5) |
| Ferritin (µg/L) | 4399 | 20.2 (19.6, 20.8) |  | 1322 | 21.3 (20.1, 22.5) |  | 1240 | 20.7 (19.5, 21.9) |  | 319 | 22.1 (19.6, 24.8) |  | 629 | 25.3 (23.6, 27.0) |  | 889 | 14.9 (14.0, 15.9) |
| Hepcidin (µg/L) | 4308 | 6.4 (6.1, 6.6) |  | 1255 | 5.6 (5.2, 6.0) |  | 1266 | 6.8 (6.3, 7.2) |  | 295 | 5.3 (4.5, 6.3) |  | 619 | 6.1 (5.5, 6.9) |  | 873 | 7.7 (7.1, 8.4) |
| sTfR (mg/L) | 4379 | 10.0 (9.8, 10.2) |  | 1344 | 18 (17.6, 18.4) |  | 1274 | 6.8 (6.5, 7.1) |  | 327 | 17.6 (16.6, 18.7) |  | 546 | 3.4 (3.3, 3.5) |  | 888 | 11.2 (10.9, 11.6) |
| Transferrin (µg/L) | 3800 | 2.7 (2.7, 2.7) |  | 1313 | 2.8 (2.7, 2.8) |  | 1282 | 2.7 (2.6, 2.7) |  | 316 | 2.7 (2.6, 2.8) |  | n/a | n/a |  | 889 | 2.7 (2.7, 2.8) |
| Iron (µmol/L) | 1664 | 6.9 (6.7, 7.0) |  | 1337 | 6.4 (6.1, 6.6) |  | n/a | n/a |  | 327 | 6.1 (5.7, 6.5) |  | n/a | 8.6 (8.3, 8.9) |  | n/a | n/a |
| TSAT (%) | 1612 | 10.1 (9.8, 10.4) |  | 1297 | 9.2 (8.8, 9.6) |  | n/a | n/a |  | 315 | 9.0 (8.3, 9.7) |  | n/a | 12.8 (12.3, 13.4) |  | n/a | n/a |
| Hemoglobin (g/dL) | 2971 | 10.4 (10.4, 10.5) |  | 793 | 10.1 (10.0, 10.2) |  | 1241 | 11 (10.9, 11.1) |  | 314 | 9.5 (9.4, 9.6) |  | 623 | 10.6 (10.4, 10.7) |  | n/a | n/a |
| Abbreviations: 25(OH)D, 25-hydroxyvitamin D; n/a, not available; sTfR, soluble transferrin receptors; TSAT, transferrin saturation; CRP, C reactive protein. Anthropometric and hemoglobin measurements were not available for the South African children. Iron was not measured in plasma samples from Ugandan and South African cohorts because they were stored in ethylenediaminetetraacetic acid (EDTA) which chelates iron. | | | | | | | | | | | | | | | | | |

**Table S3. Effect of vitamin D status on individual measures of iron status**

| **Biomarker** | ***n*** | **Overall** |  | **Vitamin D status categories** | | | | | | | | |
| --- | --- | --- | --- | --- | --- | --- | --- | --- | --- | --- | --- | --- |
|  |  |  |  | **>75 nmol/L (reference)** |  | **50–75 nmol/L** | | |  | **<50 nmol/L** | | |
|  |  | **Mean^1^ (95% CI)** |  | **Mean (95% CI)** |  | **Mean (95% CI)** | **Beta (95% CI)^2^** | ***P* value** |  | **Mean (95% CI)** | **Beta (95% CI)^1^** | ***P* value** |
| Ferritin (µg/L) | 4358 | 20.2 (19.6, 20.8) |  | 20.5 (19.6, 21.3) |  | 21.0 (20.0, 22.0) | -0.05 (-0.11, 0.01) | 0.10 |  | 15.2 (13.4, 17.2) | -0.35 (-0.45, -0.24) | <0.0001 |
| Hepcidin (µg/L) | 4265 | 6.4 (6.1, 6.6) |  | 6.4 (6.1, 6.7) |  | 6.6 (6.2, 7.1) | 0.0002 (-0.08, 0.08) | 1.0 |  | 5.3 (4.6, 6.1) | -0.30 (-0.45, -0.16) | <0.0001 |
| sTfR (mg/L) | 3791 | 10.0 (9.8, 10.2) |  | 10.6 (10.4, 11.1) |  | 9.2 (8.8, 9.5) | -0.05 (-0.08, -0.01) | 0.013 |  | 9.0 (8.3, 9.8) | -0.08 (-0.14, -0.01) | 0.029 |
| Transferrin (µg/L) | 3770 | 2.7 (2.7, 2.7) |  | 2.8 (2.8, 2.8) |  | 2.6 (2.6, 2.7) | -0.16 (-0.20, -0.12) | <0.0001 |  | 2.3 (2.2, 2.4) | -0.39 (-0.46, -0.32) | <0.0001 |
| Iron (µmol/L) | 1650 | 6.9 (6.7, 7.0) |  | 6.4 (6.2, 6.7) |  | 7.2 (7.0, 7.5) | 0.03 (-0.02, 0.09) | 0.25 |  | 8.4 (7.6, 9.2) | 0.13 (0.02, 0.23) | 0.015 |
| TSAT (%) | 1599 | 10.1 (9.8, 10.4) |  | 9.2 (8.8, 9.5) |  | 10.8 (10.3, 11.3) | 0.05 (-0.01, 0.11) | 0.082 |  | 13.5 (12.1, 15.0) | 0.21 (0.09, 0.32) | 0.0004 |
| Hemoglobin (g/dL) | 2939 | 10.4 (10.4, 10.5) |  | 10.4 (10.3, 10.4) |  | 10.5 (10.4, 10.6) | -0.04 (-0.14, 0.07) | 0.50 |  | 10.4 (10.2, 10.6) | -0.22 (-0.42, -0.03) | 0.025 |
| Abbreviations: 25(OH)D, 25-hydroxyvitamin D; sTfR, soluble transferrin receptor; TSAT, transferrin saturation; CRP, C reactive protein. **^1^**Geometric means are presented. **^2^**Beta coefficients and their *p* values were obtained from linear regression models adjusted for age, sex, season, inflammation and country. Both the dependent and independent variables (except transferrin and hemoglobin) were log-transformed before inclusion in the regression model. Thus, the beta values may be interpreted as follows for the log-transformed markers: a 1% change in 25(OH)D concentrations correspond to a beta % change in the individual iron markers. The group with 25(OH)D concentrations >75 nmol/L was the reference group in the regression analyses. | | | | | | | | | | | | |

**Table S4. Interaction between 25(OH)D concentrations and inflammation and malaria in predicting measures of iron status**

| **Biomarker** | ***n*** | **Beta (95% CI)** | ***P value*** |  | **Beta (95% CI)** | ***P value*** |
| --- | --- | --- | --- | --- | --- | --- |
| **A. Inflammation^1^** | | **Main effects (log vitamin D)** | |  | **Interaction (log vitamin D x inflammation)** | |
| Log ferritin (µg/L) | 4076 | 0.38 (0.28, 0.48) | <0.0001 |  | -0.31 (-0.52, -0.10) | 0.004 |
| Log hepcidin (µg/L) | 4019 | 0.22 (0.08, 0.36) | 0.002 |  | -0.19 (-0.47, 0.09) | 0.19 |
| Log sTfR (mg/L) | 3512 | 0.09 (0.03, 0.16) | 0.004 |  | -0.04 (-0.17, 0.08) | 0.51 |
| Transferrin (µg/L) | 3474 | 0.46 (0.39, 0.53) | <0.0001 |  | -0.07 (-0.20, 0.06) | 0.29 |
| Log iron (µmol/L) | 1359 | -0.13 (-0.23, -0.03) | 0.01 |  | 0.02 (-0.16, 0.21) | 0.82 |
| Log TSAT (%) | 1307 | -0.22 (-0.33, -0.11) | <0.0001 |  | -0.01 (-0.22, 0.20) | 0.95 |
| Hemoglobin (g/dL) | 2848 | 0.11 (-0.08, 0.29) | 0.27 |  | -0.02 (-0.39, 0.35) | 0.92 |
| **B. Malaria^2^** | | **Main effects (log vitamin D)** | |  | **Interaction (log vitamin D x malaria)** | |
| Log ferritin (µg/L) | 3187 | 0.23 (0.01, 0.35) | <0.0001 |  | 0.001 (-0.33, 0.33) | 1.00 |
| Log hepcidin (µg/L) | 3146 | -0.003 (-0.17, 0.16) | 0.98 |  | -0.08 (-0.52, 0.35) | 0.71 |
| Log sTfR (mg/L) | 2624 | 0.22 (0.14, 0.30) | <0.0001 |  | -0.04 (-0.25, 0.17) | 0.68 |
| Transferrin (µg/L) | 2585 | 0.41 (0.33, 0.49) | <0.0001 |  | 0.16 (-0.05, 0.36) | 0.13 |
| Log iron (µmol/L) | 1359 | -0.22 (-0.32, -0.11) | <0.0001 |  | 0.15 (-0.10, 0.41) | 0.24 |
| Log TSAT (%) | 1307 | -0.31 (-0.42, -0.19) | <0.0001 |  | 0.11 (-0.18, 0.39) | 0.47 |
| Hemoglobin (g/dL) | 2848 | -0.01 (-0.19, 0.17) | 0.90 |  | -0.004 (-0.47, 0.46) | 0.99 |
| Both the dependent variables (except transferrin and hemoglobin) and independent variable (25(OH)D) were log-transformed before inclusion in the linear model adjusted for age, sex, season, inflammation and study site. In analyses involving malaria, South African children were excluded because they were not exposed to malaria. ^1^Malaria parasitemia was defined as presence of *Plasmodium* parasitemia on blood film and ^2^inflammation as CRP > 5 mg/L or ACT >0.6 g/L (ACT, but not CRP was available for The Gambia). | | | | | | |

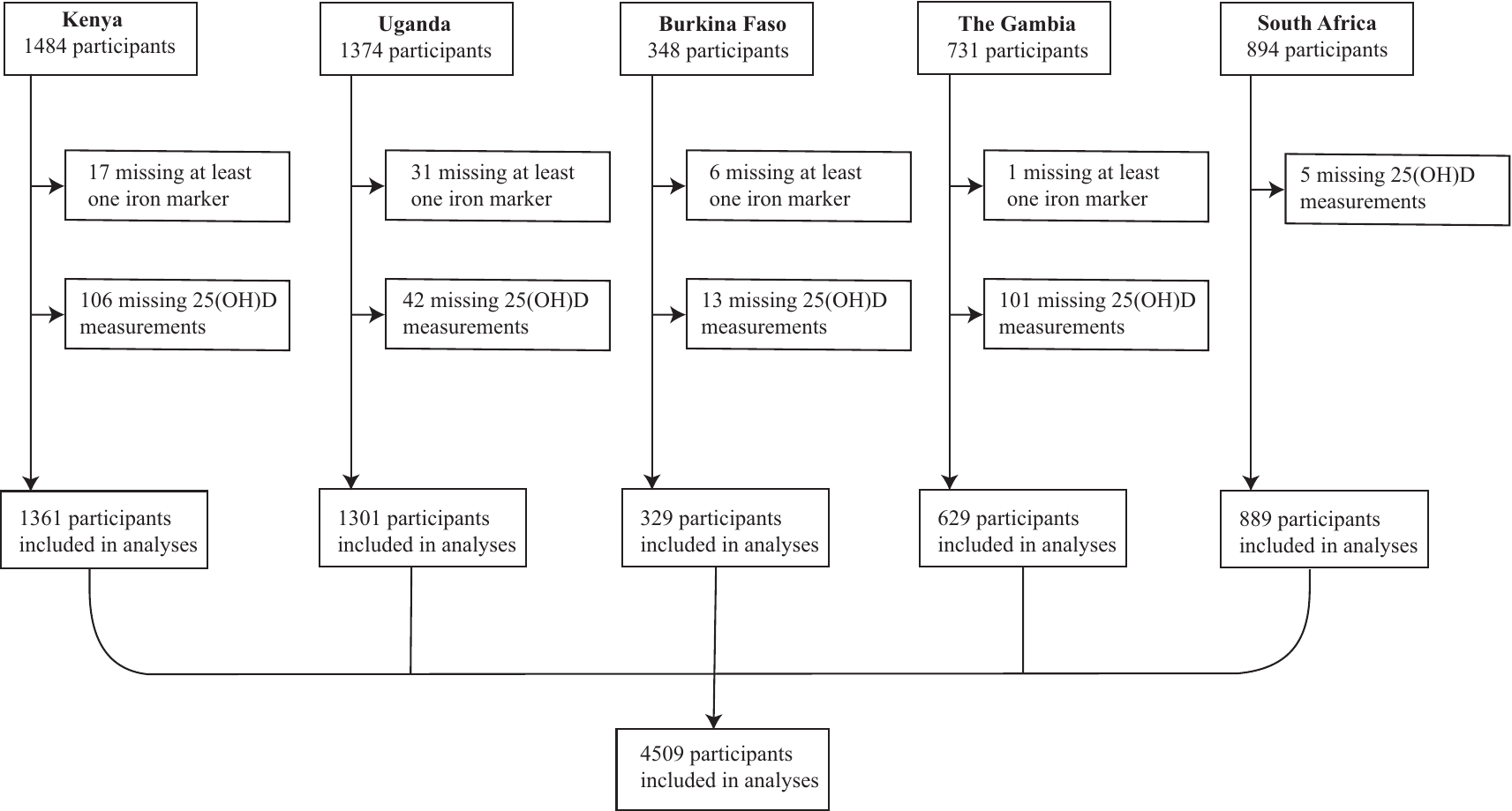

**Figure S1.** Flow chart of study participants. Iron status and 25(OH)D concentrations were measured on available stored samples from study cohorts across Africa (as described in the Methods). Study participants that did not have at least one marker of iron status or 25(OH)D concentrations measured were excluded from the analyses.

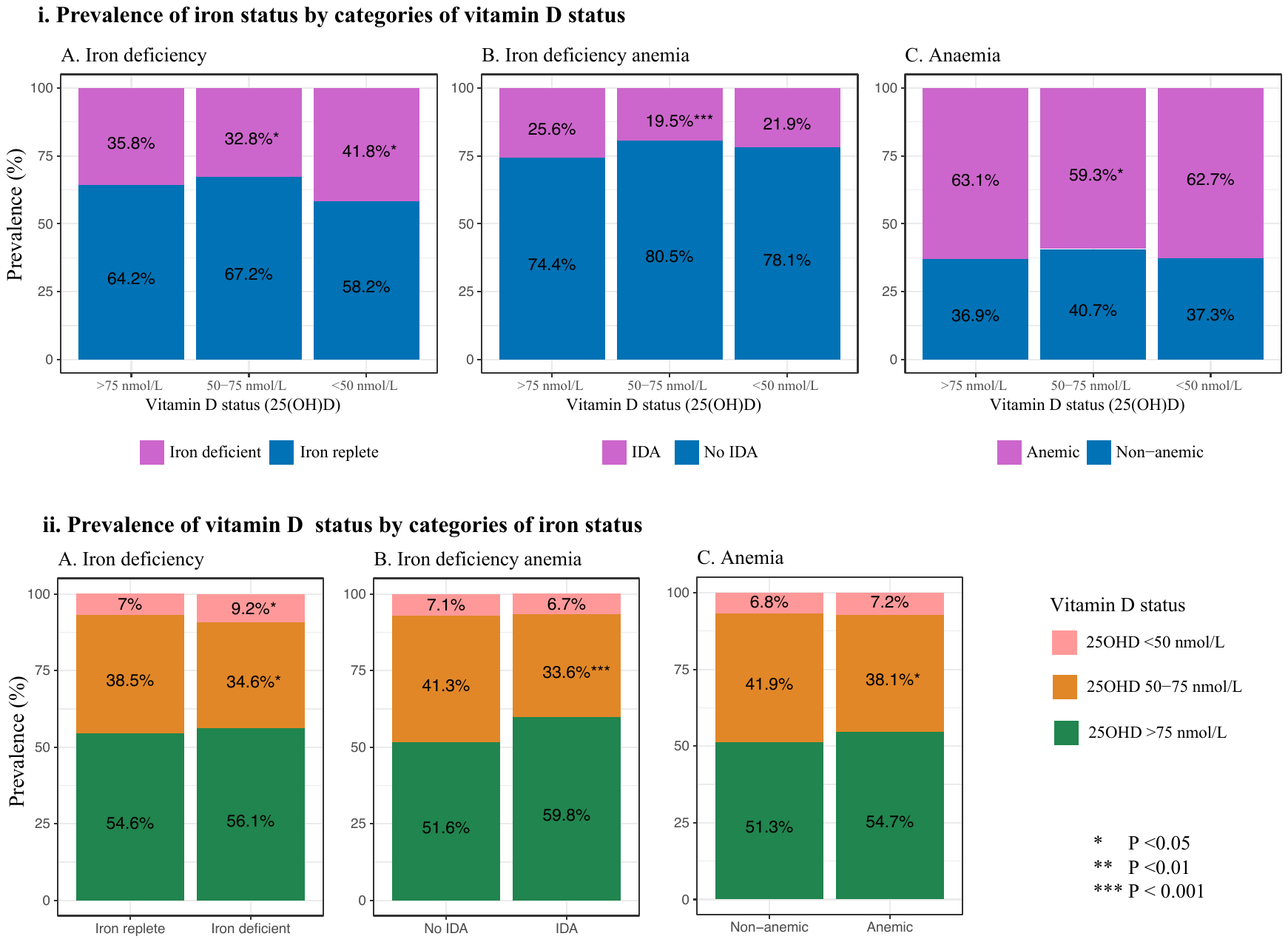

**Figure S2.** Prevalence of iron status by categories of vitamin D status (i) and prevalence of vitamin D status by categories of iron status (ii) in African children. A chi-squared test (*prtest)* was used to test the significance in the difference in proportion of low vitamin status (25(OH)D concentrations <50 or 50–75 nmol/L) within each category with the first category as the reference.

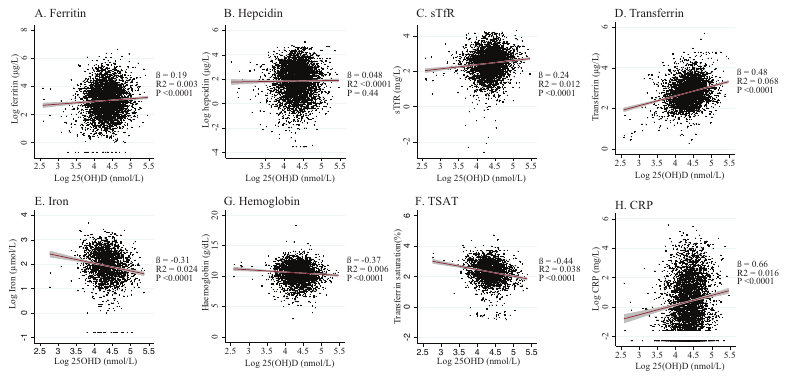

**Figure S3.** Scatter and regression plots of markers of iron status and C-reactive protein against log 25(OH)D concentrations. Abbreviations: TSAT, transferrin saturation; sTfR, soluble transferrin receptor; CRP, C-reactive protein.

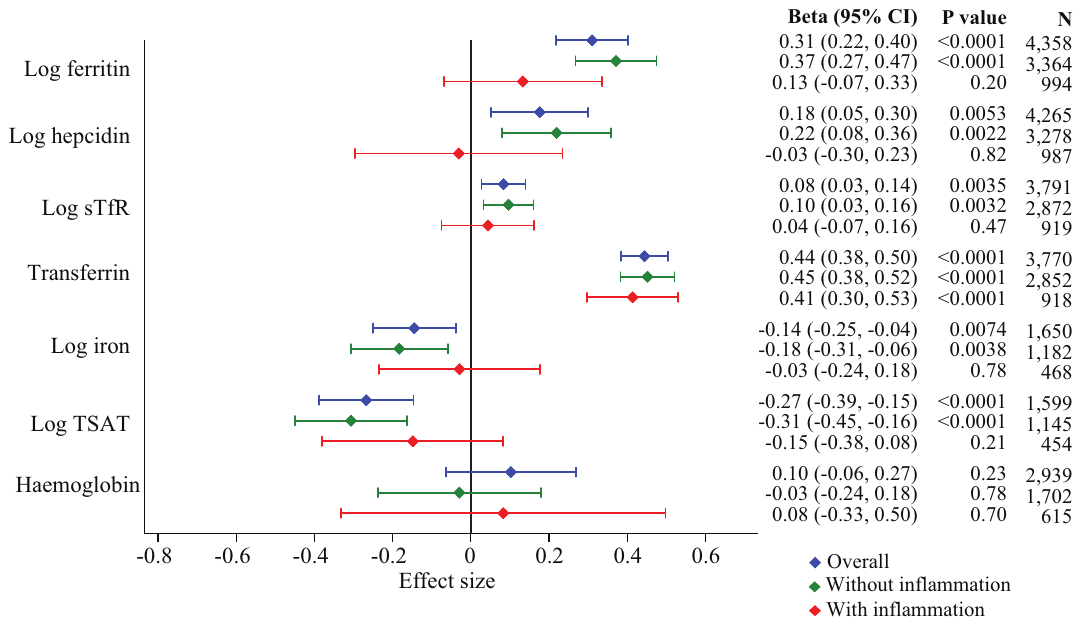

**Figure S4.** Association between 25(OH)D concentrations and iron markers overall (dark grey) and in African children with and without inflammation (black and light grey lines respectively). Both the dependent and independent variables (except transferrin and hemoglobin) were log-transformed before inclusion in the linear model. Thus the beta values may be interpreted as follows: a 1% change in 25(OH)D concentrations corresponds to a beta % change in the individual iron markers. All models were adjusted for age, sex, season, and study site and the overall model was additionally adjusted for inflammation. Error bars indicate 95% confidence intervals. Abbreviations: TSAT, transferrin saturation; sTfR, soluble transferrin receptor.

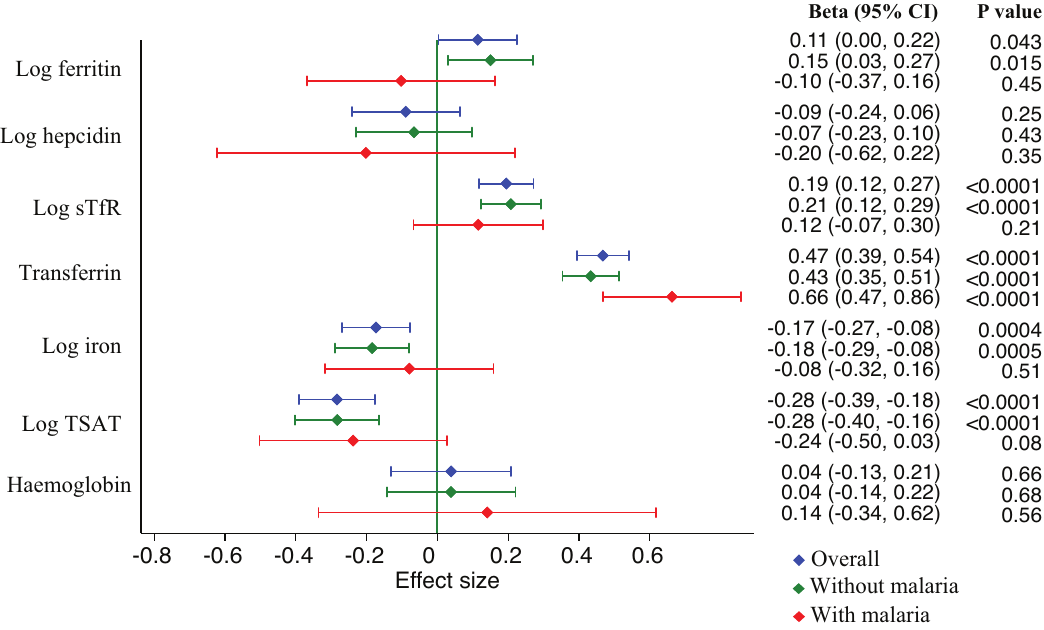

**Figure S5.** Association between 25(OH)D concentrations and iron markers overall (blue) and in children with and without malaria in Africa (red and green lines, respectively). Both the dependent and independent variables (except transferrin and hemoglobin) were log-transformed before inclusion in the linear model. Thus, the beta values may be interpreted as follows: a 1% change in 25(OH)D concentrations corresponds to a beta % change in the individual iron markers. All models were adjusted for age, sex, season, study site and inflammation and the overall model was additionally adjusted for malaria. South African children were not included in the analyses since they were not exposed to malaria. Error bars indicate 95% confidence intervals. Abbreviations: TSAT, transferrin saturation; sTfR, soluble transferrin receptor.
